# Precision therapy reduces the risk of diabetes in people with cystic fibrosis-related diabetes

**DOI:** 10.64898/2026.08.14.26360420

**Authors:** Samar E Atteih, Karen S Raraigh, Malinda Wu, Joseph M Collaco, Scott M Blackman

## Abstract

Diabetes is a highly prevalent complication of cystic fibrosis (CF), affecting 50% of adults with CF and over 80% of those with exocrine pancreatic insufficiency (PI) by age 50 years. Development of cystic fibrosis-related diabetes (CFRD) is associated with increased morbidity and mortality mostly due to advancement of chronic obstructive lung disease. Highly effective modulator therapy (HEMT), using precision medications targeting the cystic fibrosis transmembrane conductance regulator (CFTR), improves CFTR function and CF lung disease, but its impact on diabetes pathogenesis remains uncertain. We sought to determine whether two types of HEMT, ivacaftor and elexacaftor/tezacaftor/ivacaftor (ETI), alter diabetes prevalence in two large cohorts of individuals with CF and exocrine PI. For comparison, a non-highly-effective modulator, lumacaftor/ivacaftor (LUM/IVA), was also assessed. Data were provided by the CFTR2 project, a multinational CF registry (for ivacaftor and LUM-IVA), and by the CF Genome Project (CFGP), a predominantly US-based CF cohort (for ETI). Among 32,753 individuals with CF (2,803 treated), ivacaftor was associated with reduced diabetes prevalence (age-adjusted OR=0.55). In contrast, lumacaftor/ivacaftor (not highly effective) was not associated with diabetes prevalence (n=32,749). Among 2,854 individuals with CF (2,458 treated), ETI was associated with reduced diabetes prevalence (age-adjusted OR=0.47). Overall, HEMT (ivacaftor and ETI) was associated with a 25–39% reduction in diabetes prevalence in CF, while a non-highly-effective modulator (lumacaftor/ivacaftor) showed no difference. Precision targeted amelioration of CFTR dysfunction can delay onset of diabetes in a high-risk CF population.

**Highlights:**

- Diabetes is a highly prevalent complication of cystic fibrosis (CF), associated with increased morbidity and mortality. Precision medications for CF (highly effective modulator therapy, or HEMT) improve lung disease, but the impact on diabetes remains uncertain.
- We sought to determine whether HEMT use alters diabetes prevalence using two large cohorts of individuals with CF.
- Among those with similarly severe CFTR dysfunction at baseline, HEMT (ivacaftor and elexacaftor/tezacaftor/ivacaftor) was associated with a 25–39% reduction in diabetes prevalence, while non-HEMT (lumacaftor/ivacaftor) showed no difference.
- Precision targeted amelioration of CFTR dysfunction can delay onset of diabetes in a high-risk CF population.

---

Cystic fibrosis-related diabetes (CFRD) is an important age-dependent extrapulmonary complication of cystic fibrosis (CF), affecting up to 50% of adults and 20% of children.(1) CFRD occurs predominantly in those with exocrine pancreatic insufficiency (PI), a progressive process which begins in utero in those with severe CFTR dysfunction.(2) In fact, CFRD affects >80% of those with severe CFTR dysfunction by age 50 years.(3) Thus, having CF results in a predictable and very high lifetime risk of developing diabetes. As with other forms of diabetes, CFRD confers increased risks of microvascular and macrovascular complications.(4, 5)The pathogenesis of CFRD is incompletely understood but is likely multifactorial, driven in part by pancreatic beta-cell dysfunction and islet cell inflammation resulting from progressive pancreatic exocrine destruction, periductal fibrosis and global fat replacement, leading to relative insulin insufficiency.(6) This may be further compounded by insulin resistance during periods of systemic illness or prolonged glucocorticoid use.(7)

Since 2012, multiple CFTR modulators have become available to people with CF (pwCF). Some, including ivacaftor, elexacaftor/tezacaftor/ivacaftor (ETI), and vanzacaftor/tezacaftor/deutivacaftor (VTD), are considered “highly effective” based on the degree to which they restore CFTR function and improve clinical outcomes. Others, including lumacaftor/ivacaftor and tezacaftor/ivacaftor, have modest effect on CFTR function and are associated with less robust improvements in clinical outcomes, and are therefore not considered “highly effective”. Eligibility for each modulator medication is determined by an individual’s underlying CF-causing variants in CFTR which have been shown to respond. About 3% of pwCF are eligible for the first HEMT, ivacaftor, but subsequent modulator combinations were shown to be effective against more CFTR variants. Currently, over 90% of pwCF now qualify for highly effective modulator therapy (HEMT).(8)

While HEMT has dramatically transformed the CF landscape, yielding both clinical and physiologic improvements in the lung and sweat gland dysfunction, it is uncertain whether and to what extent HEMT will impact CFRD.(9) Existing evidence suggests that HEMT may be associated with reduced hemoglobin A1c,(10) reduced 2-hour oral glucose tolerance test (OGTT) glucose levels, and increased insulin secretion.(11) However, studies to date primarily address effects of HEMT on glycemic parameters and do not assess the degree to which the natural history of CFRD might be affected by HEMT use. One observational study by Volkova, et al, which included participants from both the US CF Foundation Patient Registry (CFFPR) and the UK CF Registry, showed favorable trends in the rate of increasing CFRD prevalence in ivacaftor-treated individuals, though results were not robust.(12) Furthermore, since CFRD occurs almost exclusively in those with PI, and because pancreatic fibrosis begins in utero, there has been concern that HEMT initiated after exocrine dysfunction is established may not meaningfully alter CFRD prevalence.

The primary goal of this study was to determine whether two widely used types of HEMT, ivacaftor and ETI, have impacted the prevalence of CFRD. A secondary goal was to assess the impact of a widely-used non-HEMT CFTR modulator therapy, lumacaftor/ivacaftor, on CFRD prevalence. These results could justify longitudinal analysis of the relationship between HEMT and CFRD incidence over time, to inform optimal approaches to reduce the burden of diabetes in individuals with CF.

## Methods

### Study Design and Data Collection

This is a retrospective cohort study of individuals with CF who contributed data to the CFTR2 project or the CF Genome Project.(13) CFTR2 collected data contributed primarily by national and international registries; those with data recorded on modulator use in this study were contributed by the US CFF Patient Registry(14) (CFFPR; n=33,537), the UK CF Registry (n=10,471), the European CF Society Patient Registry (ECFSPR; n=3,360), Australian Cystic Fibrosis Data Registry (ACFDR; n=776), Canadian Cystic Fibrosis Registry (n=740), Mexico (n=142), China (n=18), and Dominican Republic (n=2).

The CF Genome Project is comprised of the CF Twin and Sibling Study, the CFRD Study, the EPIC Observational study, the Gene Modifier Study, and the Gene Modifier Study of CF Liver Disease.(15) The 5 component studies collected their data at enrollment from case report forms with updated data provided from the CFFPR. All studies were approved by the institutional review boards of their institutions.

### Study Population

Cross-sectional data were collected from the most recent year of analysis (2019 for CFTR2 and 2023 for the CFGP). CF diagnosis was defined as having two pathogenic *CFTR* variants. Severe and mild CFTR dysfunction were defined based on the rate of exocrine PI reported in CFTR2. For each CFTR variant, the PI frequency was determined among individuals harboring the variant in *trans* with a known severe *CFTR* variant. Severe CFTR dysfunction was defined as a PI rate of 80% or higher, whereas mild CFTR dysfunction was defined as a PI rate of less than 50%.(16) Variants with PI frequency 50 to <80% or with fewer than 10 individuals in CFTR2 were excluded from this classification. A severe *CFTR* genotype was defined as having two severe *CFTR* variants and a mild *CFTR* genotype was defined as having one or two mild *CFTR* variants.

Age, CFRD status, and CFTR modulator use were obtained from CFTR2 and/or the CFGP study as reported to the component registries. Values for covariates (e.g., forced expiratory volume in 1 second, height, weight, liver disease) were also obtained from the registries; the most recently available values were used. HEMT use was defined as using either ivacaftor or ETI (VTD information was not included in either data set). Not using HEMT was defined as not using either ivacaftor or ETI, or missing data for ivacaftor or ETI but nonmissing data for non-highly-effective modulators (lumacaftor/ivacaftor or tezacaftor/ivacaftor). Ivacaftor when prescribed as a single agent in a genetically eligible person is an HEMT, but in people without eligible *CFTR* genotypes, ivacaftor in combination with either lumacaftor or tezacaftor is not an HEMT. For sensitivity analyses, a group with possible evidence of liver disease was defined having any of the recorded liver-related diagnoses flagged: liver cirrhosis, liver enzyme elevation, liver disease (noncirrhosis), liver steatosis, liver disease (other), acute liver failure, or complications of liver cirrhosis.

### Statistical Analysis

Cross-sectional analyses were performed separately in the CFTR2 and CF Genome Project cohorts using the most recent available data for each participant. The primary outcome was CFRD status, analyzed as a binary variable. The primary exposure was HEMT use (ivacaftor or elexacaftor/tezacaftor/ivacaftor [ETI]); lumacaftor/ivacaftor was analyzed separately as a non-HEMT comparator. Participants with type 1 or type 2 diabetes were excluded. Associations between modulator use and CFRD were evaluated using logistic regression adjusted for age. Results are reported as odds ratios (ORs) with 95% confidence intervals (CIs). Sensitivity analyses excluded individuals receiving non-HEMT modulators and examined alternative exposure definitions and subgroup analyses by markers of possible liver disease. Two-sided P values <0.05 were considered statistically significant. Analyses were performed using STATA 18 (StataCorp).

## Results

The CFTR2 project (13) contained data through 2019 with information recorded for ivacaftor (an HEMT) and lumacaftor/ivacaftor (not an HEMT), and few individuals treated with tezacaftor/ivacaftor or ETI. The subset with nonmissing age, diabetes status, and status for at least one CFTR modulator included 49,046 individuals with data collected through 2019. The primary analysis was restricted to those with severe CFTR dysfunction (“severe *CFTR* genotypes”) based on having two pathogenic variants in *CFTR* that cause exocrine PI at least 80% of the time (e.g. F508del/F508del [c.1521_1523del/c.1521_1523del], F508del/G551D [c.1521_1523del/c.1652G>A]). **Table 1** describes the primary analysis cohorts used to analyze ivacaftor (CFTR2 project) and ETI (CFGP study) treatments. **Table 2** details modulator use within the study cohorts. The CFTR2 cohort included 32,753 individuals with known ivacaftor status (yes or no), including 2,803 ivacaftor-treated individuals and 29,950 with no ivacaftor treatment. Within the 29,950 ivacaftor-untreated group were 9,808 individuals treated with a non-highly-effective CFTR modulator (all with lumacaftor/ivacaftor, none with tezacaftor/ivacaftor).

**Table 1.** *Characteristics of individuals with CF in the CFTR2 and CFGP study cohorts (severe/PI* CFTR *genotypes)*\*

|  | CFTR2 (PI) |  |  | CFGP (PI) |  |  |
| --- | --- | --- | --- | --- | --- | --- |
|  | No HEMT | HEMT | Total | No HEMT | HEMT | Total |
| **n** | 33580 | 2830 | 36410 | 867 | 2501 | 3368 |
| **Age (yr) (SD)** | 21.8 (13.1) | 23.06 (13) | 21.9 (13.1) | 32.63<br>(12.8) | 29.98<br>(10.3) | 30.66<br>(11.1) |
| **Female (%)** | 15689 (47%) | 1342 (47%) | 17031 (47%) | 418 (48%) | 1197<br>(48%) | 1615<br>(48%) |
| **Male (%)** | 17891 (53%) | 1488 (53%) | 19379 (53%) | 449 (52%) | 1304<br>(52%) | 1753<br>(52%) |
| **No liver abnormality noted** | not available | not available | not available | 572 (66%) | 1822<br>(73%) | 2394<br>(71%) |
| **Liver abnormality noted** | not available | not available | not available | 295 (34%) | 679 (27%) | 974 (29%) |
| **No CFRD** | 23949 (71%) | 2237 (79%) | 26186 (72%) | 330 (38%) | 1625<br>(65%) | 1955<br>(58%) |
| **CFRD** | 9631 (29%) | 593 (21%) | 10224 (28%) | 537 (62%) | 876 (35%) | 1413<br>(42%) |
| **Age (no CFRD) (SD)** | 18.04 (11.7) | 20.74 (12.4) | 18.28 (11.8) | 27.95<br>(10.7) | 27.75 (8.8) | 27.79 (9.1) |
| **Age (CFRD) (SD)** | 31.14 (11.6) | 31.82 (11.8) | 31.18 (11.6) | 35.5 (13.2) | 34.11<br>(11.6) | 34.64<br>(12.2) |

\* Abbreviations: SD, standard deviation; percentages are out of total for that column

**Table 2.** *CFTR modulator use in the CFTR2 and CFGP study cohorts (severe/PI* CFTR *genotypes)\**

|  | CFTR2 (PI) cohort modulator use |  |  | CFGP (PI) cohort modulator use |  |  |
| --- | --- | --- | --- | --- | --- | --- |
|  | No | Yes | Total | No | Yes | Total |
| **Ivacaftor** | 29950 (82%) | 2803 (8%) | 32753 | 3324 (99%) | 44 (1%) | 3368 |
| **Lum/Iva** | 21559 (59%) | 13244 (36%) | 34803 | 3070 (91%) | 56 (2%) | 3126 |
| **Tez/Iva** | <5 (<1%) | 453 (1%) | 494 | 2963 (88%) | 41 (1%) | 3004 |
| **ETI** | <5 (<1%) | 27 (0%) | 28 | 430 (13%) | 2458 (73%) | 2888 |
| **Any HEMT** | 33580 (92%) | 2830 (8%) | 36410 | 867 (26%) | 2501 (74%) | 3368 |
| **Any modulator** | 20142 (55%) | 16268 (45%) | 36410 | 790 (23%) | 2578 (77%) | 3368 |

\* Percentages are out of total for each cohort

The CFGP study (17) contained data through 2023 with information recorded for multiple CFTR modulators including ETI. The subset with nonmissing age, CFRD status, and status for at least one CFTR modulator included 3,963 individuals (all from the US) with data collected through 2023, including 2,888 individuals in the primary analysis cohort (severe *CFTR* genotypes, known ETI status). Data were available for 2,458 individuals treated with ETI and 430 with no ETI. The group with no ETI included 43 individuals treated with non-highly-effective CFTR modulators (18 lumacaftor/ivacaftor, 25 tezacaftor/ivacaftor).

### Ivacaftor use (CFTR2 project)

In 32,753 individuals with severe *CFTR* genotypes (mean age 21.6 years), diabetes prevalence was lower in those treated with ivacaftor (589 of 2,803, 21.0%) than in those not treated with ivacaftor (8,324 of 29,950, 27.8%; OR 0.69 [0.63-0.76], z=-7.68, p=1.6E-14). Examining the age-dependent prevalence of CFRD also revealed lower diabetes prevalence in ivacaftor-treated individuals across a range of ages (**Figure 1A**). When adjusting for age using logistic regression, the CFRD rate was again lower in the ivacaftor-treated cohort (age-adjusted OR 0.55 [0.49-0.61], n=32,753, z=-11.19, p=4.6E-29). Sensitivity analyses that excluded individuals treated with a non-highly effective CFTR modulator (lumacaftor/ivacaftor or tezacaftor/ivacaftor) again showed that ivacaftor treatment was associated with lower odds of CFRD (age-adjusted OR 0.54 [0.46-0.58], n=22,780, z=-11.64, p=2.6E-31). When including ETI use as another HEMT, use of either ivacaftor alone or ETI was also associated with lower odds of CFRD (age-adjusted OR 0.53 [0.48-0.59], n=36,410, z=-11.83, p=2.7E-32). Individuals with *CFTR* genotypes conferring residual function have retained exocrine pancreatic sufficiency and reduced risk of CFRD.(3) Our results were consistent with this concept: individuals with *CFTR* genotypes conferring residual CFTR function in CFTR2 had low risk of CFRD and no correlation between ivacaftor treatment and CFRD (age-adjusted OR=1.19 [0.97-1.5], p=0.10, n=7,370) (Supplementary Tables 1 and 2). Overall, these findings indicate that ivacaftor treatment is associated with reduced diabetes prevalence across a range of ages of people with severe *CFTR* genotypes conferring exocrine pancreatic insufficiency, which is the most common form of CF.

**Figure 1.**
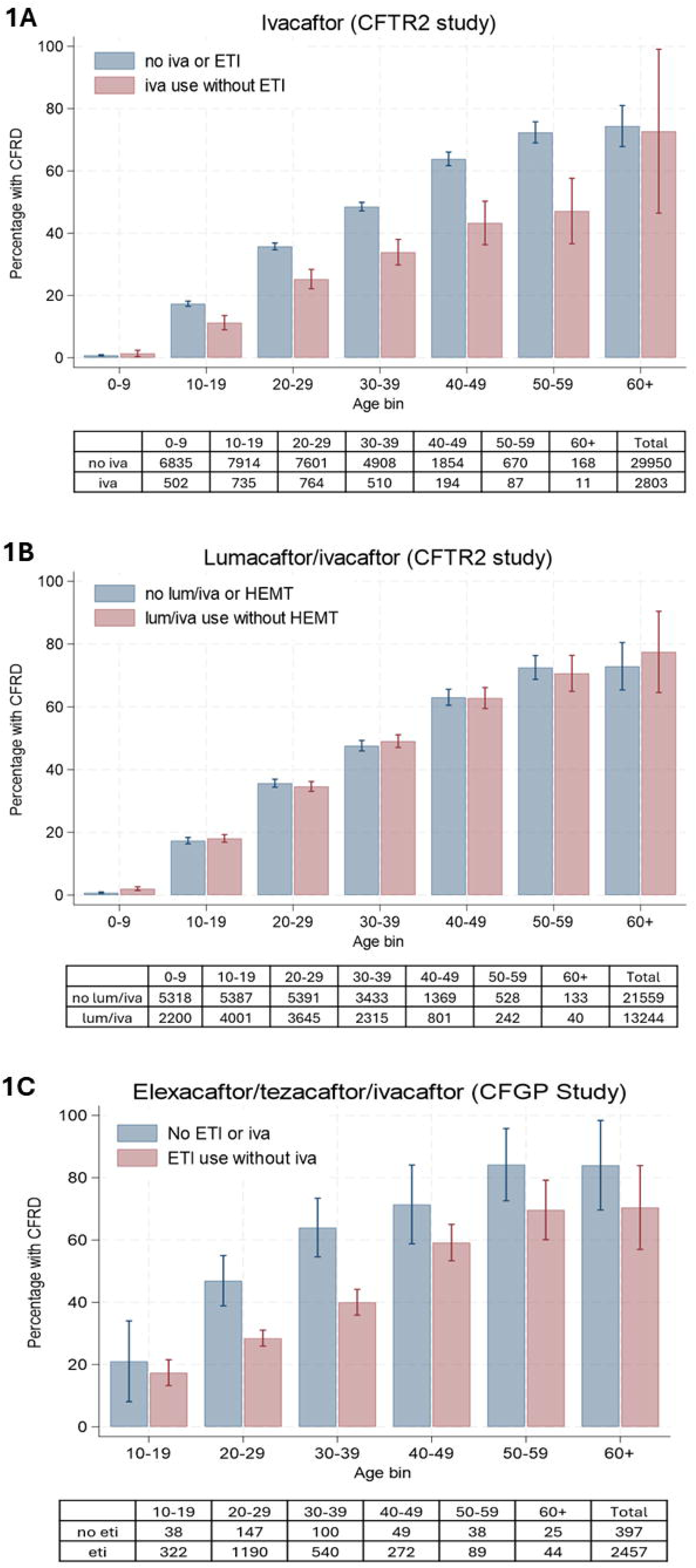
Effects of CFTR modulator terapy on risk of CFRD. Shown is the CFRD percentage by age and modulator treatment for ivacaftor (1A), lumacaftor-ivacaftor (1B), and elecacaftor/tezacaftor/ivacaftor (1C). CFRD, cystic fibrosis related diabetes; CFTR, cystic fibrosis transmembrane conductance regulator; eti, elexacaftor/tezacaftor/ivacaftor; iva, ivacaftor; lum/iva, lumacaftor/ivacaftor.

### Non-highly effective CFTR modulator use (CFTR2 project)

To account for the possibility that unmeasured biases might have contributed to the association between a medical treatment (HEMT) and an outcome (CFRD), we also considered treatment with lumacaftor/ivacaftor, which has a lower effect on CFTR function and is not considered a highly effective CFTR modulator. Nevertheless, individuals with access to ivacaftor or lumacaftor/ivacaftor might have different medical and demographic characteristics than those without such access. The CFTR2 project cohort included data from 32,749 individuals with lumacaftor/ivacaftor information (13,081 individuals treated, 19,668 not treated) who were also not recorded as having either ivacaftor or ETI treatment (**Table 2**). As shown in **Figure1B**, there was no apparent difference in the CFRD rate in those treated vs. not treated with lumacaftor/ivacaftor. Using logistic regression to adjust for age, no significant correlation was found between CFRD and lumacaftor/ivacaftor treatment (age-adjusted OR 1.02; p=0.58, n=32,749).

### ETI use (CFGP study)

In individuals with severe CFTR genotypes indicative of pancreatic insufficiency, the prevalence of diabetes was lower in those treated with ETI (865 of 2,457, 35.0%) than in those not treated with any HEMT (229 of 397, 57%; OR 0.39 [0.32-0.49], z=-8.36, p=6.3E-17). Examining the age-dependent prevalence of diabetes revealed lower prevalence in ETI-treated individuals across a range of ages (**Figure 1C**). When adjusting for age using logistic regression, the diabetes rate was again lower in the ETI-treated cohort (age-adjusted OR 0.47 [0.38-0.59], n=2,854, z=-6.41, p=1.6E-10). Sensitivity analyses revealed that ETI treatment was again associated with lower odds of CFRD (age-adjusted OR 0.47 [0.37-0.60], n=2,798, z=-6.11, p=1.0E-11) when individuals treated with any non-highly effective CFTR modulator were excluded. Use of either ivacaftor alone or ETI (both HEMT) was also associated with lower odds of CFRD (age-adjusted OR 0.34 [0.29-0.41], n=3,368, z=-12.42, p=2.0E-35). These analyses indicate this study’s findings are not dependent on exactly which form of HEMT is analyzed or whether non-highly-effective modulator treatment is included. A parallel analysis was carried out on individuals with milder CFTR dysfunction which typically confers exocrine pancreatic sufficiency (“mild *CFTR* genotypes”). When analyzing 215 individuals with mild *CFTR* genotypes, there was no statistically significant evidence for correlation between ETI treatment and CFRD (age-adjusted OR=0.20 [0.04-1.14], p=0.07, n=215) (Supplementary Tables 1 and 2). These analyses indicate that, similar to ivacaftor treatment, ETI treatment is associated with reduced diabetes prevalence across a range of ages of people with CF.

### Interaction between liver dysfunction and diabetes; effect of HEMT

Considering that CF liver disease is a risk factor for diabetes and also might affect HEMT treatment decisions, we compared a subset with a number of possible indicators of liver disease (e.g., elevated aminotransferase levels, diagnosed cirrhosis; see Methods; n=974) with those without any of the aforementioned liver disease indicators (n=2,394). There was no significant difference in ETI use between those with and without liver abnormality (OR=0.83, p=0.10, n=2,888). ETI use was correlated with reduced rate of diabetes both in the subset without evidence of liver disease (age-adjusted OR=0.50 [0.39-0.66], z=-5.02, p=5.2E-07, n=2,082) and also in the subset with evidence of liver disease (age-adjusted OR=0.62 [0.42-0.92], z=-2.39, p=0.02, n=806); no evidence was found for difference in ETI effect size depending on liver abnormality (interaction term p=0.40). Thus, HEMT use was associated with reduced diabetes risk independent from having or not having evidence of liver disease.

## Discussion

This study found that HEMT was associated with lower diabetes prevalence among pwCF with severe CFTR dysfunction across a wide range of ages, across two independent data sets, and with two distinct forms of HEMT, ivacaftor and ETI. In contrast, lumacaftor/ivacaftor, which is not considered highly effective, was not associated with reduced diabetes prevalence.

There are multiple plausible ways in which HEMT might reduce CFRD prevalence. First, it is well-established that diabetes occurs primarily in ∼85% of CF individuals with exocrine pancreatic insufficiency, as progressive exocrine dysfunction leads to pancreatic beta-cell loss and insulin insufficiency. The restoration of CFTR-mediated chloride and bicarbonate secretion in ductal epithelium may reduce ductal obstruction and resultant periductal fibrosis, fatty replacement and surrounding islet cell damage, thereby reducing pancreatic destruction.(18) Evidence supportive of this includes clinical studies showing increased fecal elastase-1 levels with ETI use, and one study from the UK showing a 28% conversion from PI to PS in preschoolers who were newly eligible for ETI.(19) Recovery of *CFTR* function *in utero* (when PI is developing) can result in preservation of exocrine pancreatic function as evidenced by case reports of infants with preserved exocrine pancreatic function whose mothers took ETI prenatally.(20, 21) When children are treated with ETI at a later age (when exocrine disease has progressed substantially)(2) the evidence is mixed, with many studies showing no significant changes in stool FE-1 levels or rate of PI to PS conversion.(22, 23) This is altogether suggestive that when initiated in early childhood or even *in utero*, HEMT use may prevent or slow the progression of pancreatic destruction, and in turn, development of CFRD. On the other hand, our study found a reduction in CFRD prevalence across a wide range of ages, suggesting that benefits may not be limited to those treated with HEMT at early ages.

A second possible mechanism is that HEMT may delay progression of endocrine dysfunction, or perhaps improve existing endocrine dysfunction. Whether this is the case remains unclear, with current evidence mixed but less supportive of functional recovery of already damaged beta-cells. CFTR expression has been identified in beta-cells, where it may modulate insulin secretion directly, though this has been debated. Some studies have suggested that CFTR may instead influence beta-cell function indirectly via paracrine signaling from ductal cells. In CFTR loss-of-function models, centroacinar cells, which are progenitor cells contributing to beta-cell regeneration, have been found to be deficient.(24) Therefore, it is possible that impaired centroacinar cell renewal contributes to progressive beta-cell loss. If so, it is possible that the restoration of CFTR function via HEMT could preserve progenitor cell populations and slow progressive beta-cell loss. The PROMISE-ENDO prospective multicenter study found no significant change in insulin secretion at 24-30 months following ETI initiation, though there were modest improvement in HbA1c and fasting glucose levels.(11) Another study demonstrated that amongst pwCF with pre-existing impaired glucose tolerance or CFRD, ETI use led to improved OGTT results, with reduction in mean 2 hour glucose levels and improved dysglycemia categorization over 48 weeks.(25) This suggests that some degree of functional recovery is possible. This cross-sectional study was not designed to test for recovery of CFRD, but longitudinal analyses will be sensitive to this possibility.

Third, it is possible that HEMT leads to indirect benefits in CFRD development. While CFRD is primarily caused by insulin insufficiency, periods of increased insulin resistance and hyperglycemia may occur in setting of recurrent exacerbations and systemic inflammation, or during oral corticosteroid use for treatment of complications such as allergic bronchopulmonary aspergillosis.(18) HEMT may decrease the risk of CFRD indirectly by reducing the frequency of pulmonary exacerbations, systemic inflammation and systemic steroid use which may all contribute to the progression of glycemic abnormalities.(18, 26, 27)

CFTR modulator therapy has evolved substantially since ivacaftor was first approved in 2012 as a single-agent potentiator for pwCF with gating variants (harbored by approximately 5% of pwCF). Ivacaftor increases the open probability of correctly folded CFTR already on the cell surface and is therefore highly effective for gating (Class III) and select residual function variants (Class IV-V), in which variable quantities of functionally normal CFTR protein are produced and trafficked to the cell surface. It is, however, ineffective for Class II variants, such as F508del, in which the CFTR protein is misfolded and fails to traffic to the cell surface.(28) The addition of the first-generation corrector lumacaftor to ivacaftor in 2015 extended modulator eligibility to those homozygous for F508del, but lumacaftor only partially rescues folding and trafficking impairment, so ivacaftor is potentiating only partially restored protein at the cell surface and yields only modest clinical benefit.(29) Tezacaftor/ivacaftor followed in 2018 with improved tolerability but again limited efficacy, and eligibility restricted to F508del homozygotes and select residual function variants.(30) The approval of elexacaftor/tezacaftor/ivacaftor (ETI) in 2019 marked a transformative advance in CF care. ETI pairs ivacaftor with two next-generation correctors that achieve far greater CFTR rescue for pwCF with at least one F508del allele (approximately 90% of the CF population) and has been shown to be highly effective.(31)

The absence of association between lumacaftor/ivacaftor and CFRD prevalence is consistent with its modest effect on CFTR function and with prior work showing that lumacaftor/ivacaftor does not improve insulin secretion.(32) This is a notable finding because it suggests that a certain threshold of CFTR restoration may be necessary to improve diabetes prevalence, even in large population analyses. It is also possible that lesser degrees of CFTR restoration confer slower and more subtle improvement in diabetes risk, and perhaps our observation period for lumacaftor/ivacaftor was insufficient to observe these changes.

Our study has several strengths, and builds on prior work by Volkova et al., which showed favorable trends in CFRD prevalence amongst pwCF treated with ivacaftor across both US and UK CF patient registries over the five years following its commercial availability.(12) While the prior analysis was limited to ivacaftor-treated individuals with gating variants, our use of two independent, large-scale datasets which encompassed both national and international registry data spanning multiple countries, provided a larger and more geographically diverse sample. We additionally demonstrated an association between HEMT and reduced CFRD prevalence not only with ivacaftor but also with ETI, which targets a far broader population of pwCF with severe CFTR dysfunction. This association was strengthened by our inclusion of lumacaftor/ivacaftor (non-HEMT) as comparator group, which was available to a similar population to ETI and across an overlapping time frame. Since there was no observed association between lumacaftor/ivacaftor and diabetes prevalence, this suggests that findings are not primarily driven by confounding factors, such as access to medications or healthcare utilization. Our stratification by severe CFTR dysfunction is also essential to make appropriate comparisons between treated and untreated individuals, because HEMT eligibility also depends on *CFTR* genotype. Excluding HEMT-eligible individuals with milder *CFTR* dysfunction helps reduce this type of potential confounding. Additionally, the ivacaftor and lumacaftor/ivacaftor analyses (CFTR2, data through 2019) included only data collected before the COVID-19 pandemic onset and in-person restriction implementation (March 2020). This is significant because both in-person CF clinic encounters and as a result, diabetes screening, declined substantially after pandemic onset in March 2020.(33) Because the COVID-19 pandemic coincided with the approval and beginning of widespread uptake of ETI (October 2019), there has been concern that any observed reduction in diabetes prevalence with ETI use may be confounded by decreased screening and diagnoses during this period. However, in our study, the association between HEMT and reduced diabetes prevalence was demonstrated both pre-pandemic, with ivacaftor, and during the pandemic, with ETI. This is reassuring that the observed reduction in diabetes prevalence associated with ETI is not driven solely by reduced screening and healthcare access during the pandemic. Lastly, in the CFGP subset we found that having a diagnosis of liver disease (a potential confounder) did not correlate with HEMT use, nor was there an interaction with the correlation between HEMT use and diabetes.

There are also important limitations to acknowledge. This was a cross-sectional study, and therefore we cannot establish temporal relationships or fully elucidate how HEMT impacts the natural history of CFRD. Almost all of the study participants had severe *CFTR* dysfunction, a risk factor for both exocrine PI and CFRD. We had reduced power for detecting an effect of HEMT on CFRD risk in those with milder *CFTR* genotypes. Lastly, the study was limited by availability of registry-level data in both analysis cohorts. This includes CFRD being defined by clinical diagnosis in the CFTR2 and CFGP studies, with detailed information about glucose metabolism not able to be included. In addition, information about modulator treatment starting age, duration, adherence, and discontinuation were not included. Duration of HEMT use will be particularly important to account for in future studies, as longer exposure period may lead to more pronounced reduction in CFRD risk with time.

Future directions will include longitudinal analyses to assess the relationship between HEMT and CFRD risk modification, accounting for age at modulator initiation, degree of exocrine or endocrine pancreatic dysfunction at initiation, duration of modulator treatment, and effects of modulator discontinuation. In conclusion, our cross-sectional data suggests that HEMT is associated with reduced prevalence of CFRD in pwCF with severe CFTR dysfunction, while non-HEMT is not. Our findings support the hypothesis that sufficient restoration of CFTR function, even in adulthood, modifies the trajectory of diabetes in individuals with CF. Having CF confers a very high lifetime risk of diabetes, but delay or prevention of diabetes in turn means reduced morbidity and improved longevity to individuals with CF.

## Data Availability

Summary data produced in the present study are available upon reasonable request to the authors. Individual-level data are available from the contributing registries.

## Acknowledgements

The authors would like to thank the Cystic Fibrosis Foundation for the use of CF Foundation Patient Registry data to conduct this study. Additionally, we would like to thank the patients, care providers, and clinic coordinators at CF Centers throughout the United States for their contributions to the CF Foundation Patient Registry.

## Author contributions

S.M.B. conceived and designed the study, performed statistical analysis and contributed to the interpretation of the data and revision of the manuscript, has full access to all the data in the study, and takes responsibility for the integrity of the data and the accuracy of the data analysis. K.S.R. contributed to data collection, statistical analysis, data interpretation, and manuscript revisions. J.M.C. contributed to statistical analyses, data interpretation, and manuscript revisions. M.W. Contributed to data interpretation and manuscript revisions. S.E.A. has full access to all the data in the study, contributed to statistical analyses, interpretation of the data, and wrote and revised the manuscript. All authors read and approved the final manuscript.

## Abbreviations

ACFDR: Australian Cystic Fibrosis Data Registry
CF: cystic fibrosis
CFFPR: Cystic Fibrosis Foundation Patient Registry
CFGP: CF Genome Project
CFRD: cystic fibrosis-related diabetes
CFTR: cystic fibrosis transmembrane conductance regulator
CFTR2: Clinical and Functional Translation of CFTR
CI: confidence interval
ECFSPR: European Cystic Fibrosis Society Patient Registry
ETI: elexacaftor/tezacaftor/ivacaftor
FE-1: fecal elastase-1
FEV1: forced expiratory volume in 1 second
HbA1c: hemoglobin A1c
HEMT: highly effective modulator therapy
OGTT: oral glucose tolerance test
OR: odds ratio
PI: pancreatic insufficiency
PS: pancreatic sufficiency
pwCF: people with cystic fibrosis
VTD: vanzacaftor/tezacaftor/deutivacaftor

