## Supplemental Tables 1 and 2 for "Precision therapy reduces the risk of diabetes in people with cystic fibrosis-related diabetes"

**Supplementary Table 1**

Title: *Characteristics of individuals with CF in the CFTR2 and CFGP study cohorts (mild/PS* CFTR *genotypes)**

|  | **CFTR2 (PS)** | | | **CFGP (PS)** | | |
| --- | --- | --- | --- | --- | --- | --- |
|  | **No HEMT** | **HEMT** | **Total** | **No HEMT** | **HEMT** | **Total** |
| **N** | 4908 | 2541 | 7449 | 111 | 166 | 277 |
| **Age (yr) (SD)** | 27.4 (19.2) | 29.65 (20.6) | 28.17 (19.7) | 26.42 (13.6) | 30.49 (12.5) | 28.86 (13.1) |
| **Female (%)** | 2529 (52%) | 1314 (52%) | 3843 (52%) | 44 (40%) | 98 (59%) | 142 (51%) |
| **Male (%)** | 2379 (48%) | 1227 (48%) | 3606 (48%) | 67 (60%) | 68 (41%) | 135 (49%) |
| **No liver abnormality noted** | not available | not available | not available | 105 (95%) | 155 (93%) | 260 (94%) |
| **Liver abnormality noted** | not available | not available | not available | 6 (5%) | 11 (7%) | 17 (6%) |
| **No CFRD** | 4641 (95%) | 2356 (93%) | 6997 (94%) | 105 (95%) | 163 (98%) | 268 (97%) |
| **CFRD** | 267 (5%) | 185 (7%) | 452 (6%) | 6 (5%) | <5 (<3%) | 9 (3%) |
| **Age (no CFRD) (SD)** | 26.22 (18.7) | 27.89 (19.9) | 26.78 (19.1) | 26.12 (13.7) | 30.33 (12.5) | 28.68 (13.1) |
| **Age (CFRD) (SD)** | 48.01 (15.4) | 52.11 (14.6) | 49.69 (49.7) | 31.63 (11.2) | 39.2 (13.8) | 34.16 (11.9) |

* Abbreviations: SD, standard deviation; percentages are out of total for that column

**Supplementary Table 2**

Title: *CFTR modulator use in the CFTR2 and CFGP study cohorts (mild/PS* CFTR *genotypes)**

|  | **CFTR2 (PS) cohort modulator use** | | | **CFGP (PS) cohort modulator use** | | |
| --- | --- | --- | --- | --- | --- | --- |
|  | **No** | **Yes** | **Total** | **No** | **Yes** | **Total** |
| **Ivacaftor** | 4830 (65%) | 2540 (34%) | 7370 | 246 (89%) | 31 (11%) | 277 |
| **Lum/Iva** | 6834 (92%) | 88 (1%) | 6922 | 261 (94%) | <5 (<2%) | 261 |
| **Tez/Iva** | 11 (0%) | 60 (1%) | 71 | 237 (86%) | 18 (6%) | 255 |
| **ETI** | 11 (0%) | <5 (<1%) | 12 | 106 (38%) | 136 (49%) | 242 |
| **Any HEMT** | 4908 (66%) | 2541 (34%) | 7449 | 111 (40%) | 166 (60%) | 277 |
| **Any modulator** | 4810 (65%) | 2639 (35%) | 7449 | 97 (35%) | 180 (65%) | 277 |

* Percentages are out of total for each cohort
